# Mitigating outbreaks in congregate settings by decreasing the size of the susceptible population

**DOI:** 10.1101/2021.07.05.21260043

**Authors:** Seth Blumberg, Phoebe Lu, Christopher M. Hoover, James O. Lloyd-Smith, Ada T. Kwan, David Sears, Stefano M. Bertozzi, Lee Worden

## Abstract

While many transmission models have been developed for community spread of respiratory pathogens, less attention has been given to modeling the interdependence of disease introduction and spread seen in congregate settings, such as prisons or nursing homes. As demonstrated by the explosive outbreaks of COVID-19 seen in congregate settings, the need for effective outbreak prevention and mitigation strategies for these settings is critical. Here we consider how interventions that decrease the size of the susceptible populations, such as vaccination or depopulation, impact the expected number of infections due to outbreaks. Introduction of disease into the resident population from the community is modeled as a branching process, while spread between residents is modeled via a compartmental model. Control is modeled as a proportional decrease in both the number of susceptible residents and the reproduction number. We find that vaccination or depopulation can have a greater than linear effect on anticipated infections. For example, assuming a reproduction number of 3.0 for density-dependent COVID-19 transmission, we find that reducing the size of the susceptible population by 20% reduced overall disease burden by 47%. We highlight the California state prison system as an example for how these findings provide a quantitative framework for implementing infection control in congregate settings. Additional applications of our modeling framework include optimizing the distribution of residents into independent residential units, and comparison of preemptive versus reactive vaccination strategies.

## 1 Introduction

The COVID-19 pandemic has highlighted the need to quickly identify and implement strategies for controlling the spread of a novel respiratory pathogen. A particular challenge in congregate settings such as prisons, nursing homes, and crowded workplaces where transmission is amplified^1–6^. The increased risk of transmission in these settings results in a higher potential for an outbreak to cause many infections within a few weeks. In addition, residents of congregate settings often have a higher prevalence of comorbidities that contribute to worse disease outcomes^7,8^. The subsequent surge of hospital admissions can strain healthcare capacity and seed increased transmission within the wider community^9,10^. Outbreaks in prison settings are further complicated by the additional security, training, and contractual resources needed to hospitalize an incarcerated person. One opportunity to reduce the public health risk associated with congregate settings is simply to decrease the number of susceptible individuals. This might occur via vaccination or depopulation^10,11^. Reducing the number of susceptible individuals can both decrease the chance of an outbreak occurring and the size of any outbreaks that occur^12^. To provide a quantitative framework to evaluate the impact of reducing the susceptible population, we describe a model for the probability of an outbreak occurring in a congregate setting within a specified time period, as well as the size of an outbreak that may occur. To illustrate the applicability of our model, we utilize publicly available data from the California Department of Corrections and Rehabilitation (CDCR) for the spread of SARS-CoV-2.

## 2 Methods

### 2.1 Data

COVID-19 data for all 35 California state prisons operated by the CDCR are reported daily in a public data dashboard. Machine readable time series of these daily reports were acquired from the University of California Los Angeles COVID Behind Bars project which gathers and organizes COVID-19 data from carceral institutions across the United States.^13^ Time series of incident cases were derived by taking the daily difference of reported cumulative cases. Differences in daily cumulative case counts that resulted in negative incidence estimates were ignored and incidence was estimated from the next reported cumulative case count that did not result in a negative incidence estimate.

### 2.2 Model overview

We structure outbreak dynamics in congregate settings into three stages (Figure 1). First, a case has to be introduced into the congregate setting. This may occur due to direct transfer of an infected case into the congregate population, or from transmission from a staff worker or visitor. We model the primary mode of introduction as being from staff introductions, since this is the hardest type of introduction to repress. Second, an introduction of a case (or a few cases) can either be self-limited or progress to a full outbreak. The probability of an outbreak occurring is impacted by the stochastic nature of disease spread, the reproduction number, and the degree of transmission heterogeneity. The reproduction number, *R*_0_, is the average number of cases each new case causes when all of their contacts are susceptible to disease. We use the dispersion parameter, *k*, to denote the amount of transmission heterogeneity. Third, if an outbreak is established, the overall impact is in proportion to the total number of cases infected. Since the large number of cases overwhelms the stochasticity of transmission, this third stage is deterministic in nature. We employ a deterministic susceptible-exposed-infectious-recovered (SEIR) compartmental model for this stage. Assumptions of the SEIR model include that the duration of natural or acquired immunity is long enough so that re-infections are unlikely within the time frame of a single outbreak.

**Figure 1.**
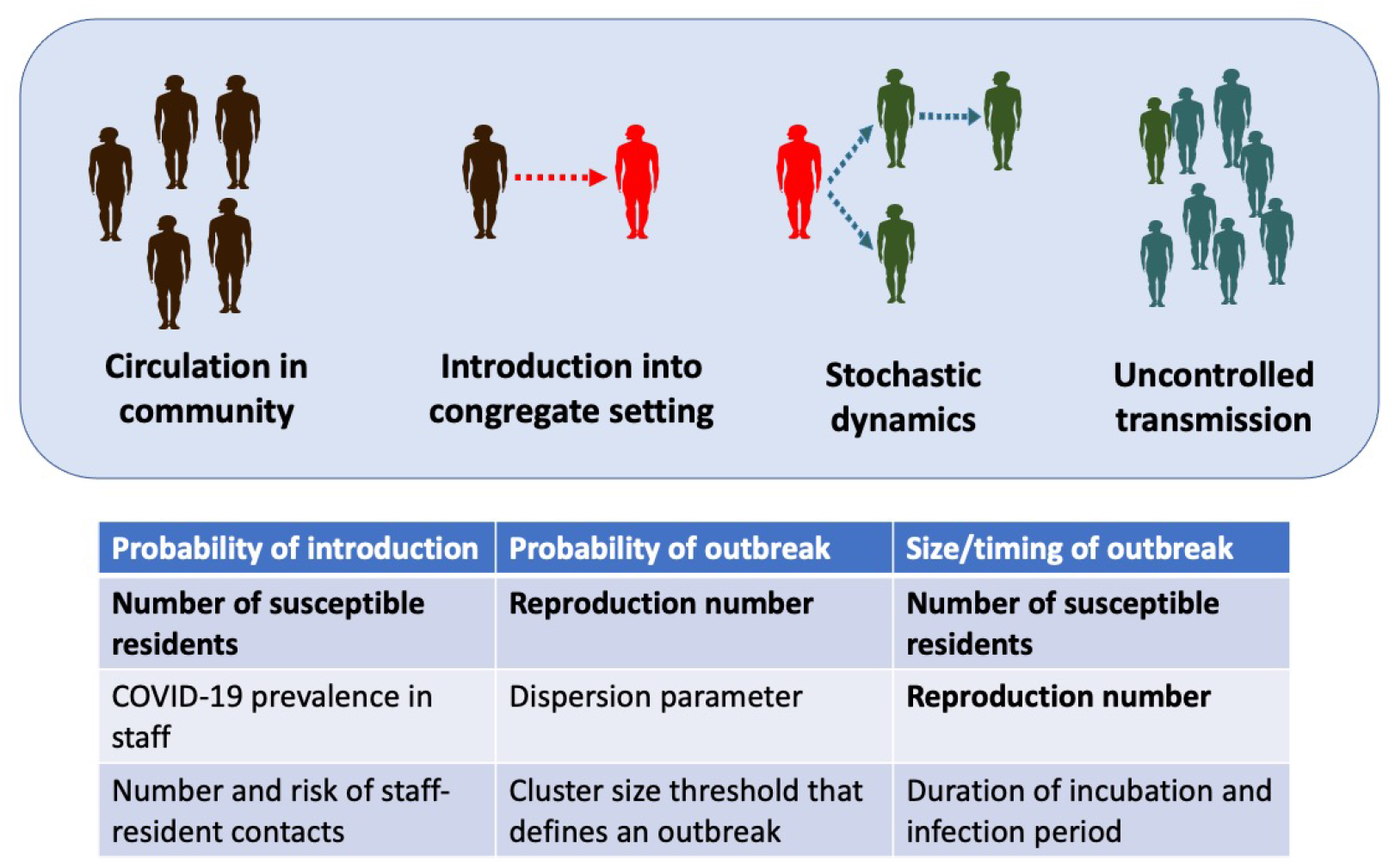
Stages of an outbreak. For an outbreak to occur in a congregate setting, an infection that circulates in the community must be introduced into the congregate setting. Then stochastic dynamics determine whether or not an introduction initiates a large outbreak. Once an outbreak occurs, uncontrolled transmission dictates the size and time course of the outbreak. The table lists the variables that are used in each step of the model. The bolded variables are impacted by a reduction in the number of susceptible individuals, as occurs with vaccination or depopulation.

We assume that *R*_0_ > 1, because when *R*_0_ is less than one, transmission is self-limited and outbreaks that involve a large portion of the resident population are not expected. The key outputs for the three stages of the model are the rate of introductions, *ϕ*, the probability that an introduction results in uncontrolled transmission, *P*_*uc*_, and the expected size of any outbreaks that occur, *I*_*tot*_. Our models for each stage of an outbreak are described in the supplementary methods.

### 2.3 Parameterization

The value of the reproduction number is of particular importance for determining the relative impact of control interventions. For any one disease, there is often great variability of reproduction number estimates. For SARS-CoV-2, there are published estimates of the reproduction number as high as 8.4 in prison settings.^14^ Meanwhile less transmissible diseases can also cause outbreaks in congregate settings, such as tuberculosis in prisons or influenza in nursing homes.^15–19^ Given the range of estimates for the reproduction number of any single disease, including variants of concern, we explore a range of values for the reproduction number. For the purpose of our analyses, *R*_0_ is defined within the context of system-wide control interventions that are in place before consideration of vaccination or depopulation. That is *R*_0_ may incorporate interventions such as masking, social distancing, ventilation improvements, improved hygiene and other practices that do not specifically reduce the number of susceptible residents. Based on current literature, we assume a latent period of three days (i.e. time between the occurrence of infection and the onset of infectiousness), and an infectious period of seven days.^20–24^

Our analyses focus mostly on the relative rather than absolute impact of reducing the size of susceptible populations. As such the specific size of the modeled populations are of secondary importance. However, to provide context for our analyses, we continue to focus on the spread of SARS-CoV-2 within the California prison system for choosing the remaining parameters. The average population size of the 35 CDCR prisons was about 3,300 at the beginning of 2020.^25^ Meanwhile prisons typically consist of multiple buildings, that have a degree of independence. Thus, to approximate a single congregate population, we choose a population size of 1,000 for our analyses. The prevalence of disease in the staff, the average daily number of staff contacts each resident has, and the probability that an infected staff transmits disease to a resident during a contact were estimated as 0.01%, 10 and 1% respectively based on a combination of community dynamics and the empirical observation of relatively frequent outbreaks occurring in the CDCR institutions^13^.

Numerous studies have shown that infectious diseases tend to exhibit superspreading behavior characterized by a low dispersion parameter^26^. Recent studies indicate that SARS-CoV-2 follow this pattern and henceforth we assume a value of 0.2 for the dispersion parameter.^27–29^.

### 2.4 Impact of decreasing the susceptible population by vaccination or depopulation

Each of the stages of our model for outbreak dynamics is influenced by either the number of susceptible individuals, *R*_0_, or both. This serves as the basis for assessing how the reduction of the susceptible population by vaccination or depopulation can impact the burden of disease in congregate settings.

We define *N*_0_, and *R*_0_ as the size of the congregate population and the reproduction number prior to any control interventions that decrease the number of susceptibles, and prior to any outbreak occurring. We let *γ* represent the amount of ‘control’ as determined by the proportional reduction in the size of the susceptible population that occurs with vaccination and/or depopulation,

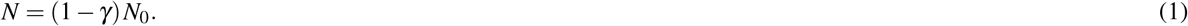

We assume a linear relationship between the effective reproduction number *R*, and *γ*,

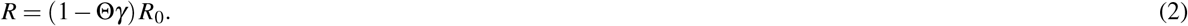

Here we have defined a susceptibility index, Θ. For Θ = 0, the reproduction number remains constant even when the number of susceptible residents are reduced. This would be equivalent to a vaccine that eliminates disease but has no impact on transmission. In the case of depopulation, Θ = 0 would be consistent with the mean number of contacts per resident remaining constant even when the population size is reduced. In contrast when Θ = 1, the ratio between the size of the susceptible population and the reproduction number remains constant. This corresponds to a vaccine that is equally effective at reducing disease and transmission in a well-mixed population. In the case of depopulation Θ = 1 corresponds to resident contacts being reduced in proportion to the population size. In disease dynamics literature Θ = 0 and Θ = 1 may be referred to as density-independent and density-dependent transmission respectively. We assume that the dispersion parameter is independent of *γ*. Intermediate values of the susceptibility index (ie. 0 *<* Θ *<* 1) may occur in many ways. Examples include vaccines that protect against disease more than against infection, or when poor ventilation implies that halving the number of infected neighbors does not half the risk of acquiring disease.

To model the overall impact of depopulation, we define *E* to be the average number of cases expected due to outbreaks that are initiated over a defined time interval, *T*. The expected number of introductions is *ϕ T*. The overall probability, *P*_*ob*_ of an outbreak occurring is one minus the probability that no introductions lead to uncontrolled transmission. The average number of cases expected due to outbreak dynamics is the overall probability that an outbreak occurs times the expected size of an outbreak. That is,

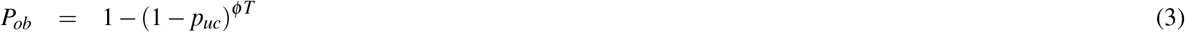

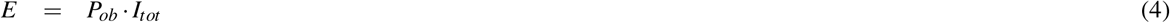

Here we have assumed that only one uncontrolled outbreak can occur in a residential unit because enough individuals would be infected so that there would be subsequent herd immunity. Each of *ϕ, P*_*ob*_, and *I*_*tot*_ depend on *γ*. The overall impact of control is probed by evaluating how *E* depends on *γ*.

In the supplemental methods, we extend this model to consider two decision-making scenarios. First, we consider how the average number of infections from outbreaks depends on how a fixed number of susceptible residents are distributed into two independent residential units. Second, in contrast to the preemptive strategy of decreasing the susceptible population before an outbreak occurs, we consider how the average number of infection from outbreaks is impacted by a reactive control strategy in which the susceptible population is rapidly reduced once an outbreak is known to occur.

All calculations and simulations are conducted in R, version 4.0.2. Code is available on github (https://github.com/proctor-ucsf/Transmission-in-congregate-settings).

## 3 Results

### 3.1 Evaluating model assumptions

From the beginning of CDCR reporting in April of 2020 through February 2, 2021 there were over 41,000 SARS-CoV2 cases in California State prisons. Within each institution, periods of low disease prevalence were interrupted by focal outbreaks caused by rapid spread (Figure 2). Some institutions have multiple outbreaks, which is consistent with separate residential buildings on a campus having outbreaks at different times. Meanwhile there are many examples of small clusters of cases that do not progress to an outbreak. Upon aggregating case counts by institution and then assigning clusters of infection based on having at least fourteen days of no cases between clusters, we find that 23% of clusters consist of an isolated case and 53% of clusters have greater than ten cases suggestive of a large outbreak. Overall this data supports our key assumptions that outbreak dynamics are supported by sporadic introductions into the residential community and that stochastic dynamics determine the probability that a sporadic introduction progresses into an uncontrolled outbreak.

**Figure 2.**
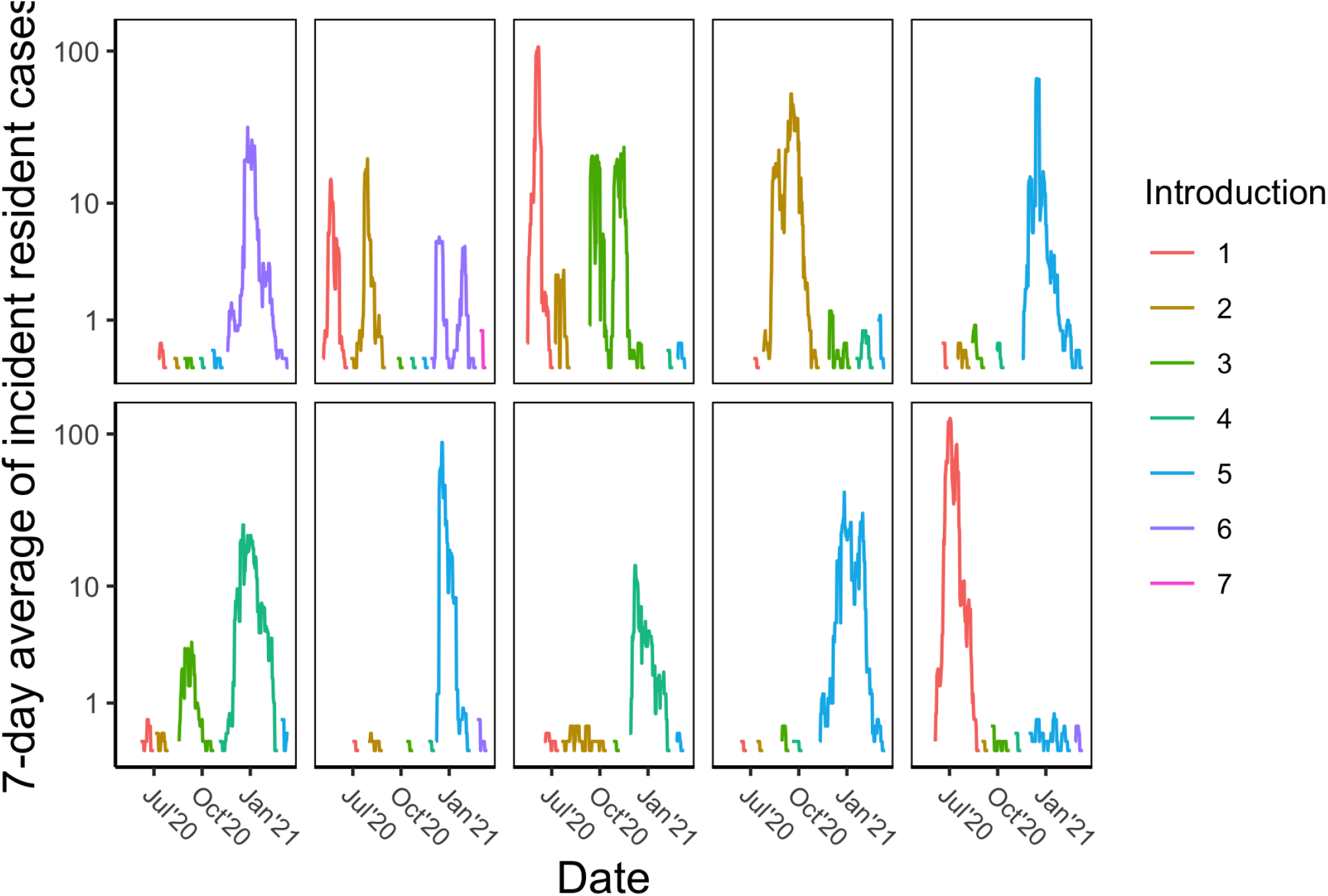
COVID-19 incidence in California State prisons. Each panel represents one state institution. For visualization purposes, Y axes are log transformed and 7-day rolling averages of incident counts are displayed. To highlight the stochastic impact of disease introductions, a new color is used for the incidence data whenever there is a period of no cases lasting at least 14 days. Thus the different colors approximate the consequence of individual disease introductions into the residential community. The panels represent all institutions where at least five introductions have occurred. Names of state prisons have been removed, but are available by request.

### 3.2 Stages of outbreak dynamics

A prerequisite for an outbreak to occur is introduction of disease into the residential community. For our model, the frequency of introductions increases with a larger residential population, increased prevalence of disease in the community, a higher resident-staff contact rate, or higher probability that a resident contact with an infected staff causes disease (Figure S1).

A disease introduction may or may not lead to an outbreak. The probability of an outbreak increases as the number of introductions, or the reproduction number increases (Figure S2). This probability also depends on the dispersion parameter. High values of the dispersion parameter correspond to homogeneous transmission and more predictable dynamics, whereas low values correspond to heterogeneous dynamics that are more likely to produce either explosive outbreaks or dead-ends to transmission. Thus high values of the dispersion parameter lead to a higher outbreak probability (Figure S2).

Once an outbreak occurs, the number of infectious individuals in our model grows exponentially until there is a significant depletion of susceptible individuals (Figure S3). A reproduction number above one is necessary for an outbreak to occur, but even values moderately above one lead to a large attack rate (i.e.proportion of infected residents). A reproduction number as low as 1.5 results in an attack rate of 59%, and a reproduction number of 2.5 leads to an attack rate of 89% (as seen by the asymptotic value for the number of removed individuals in the two panels of Figure S3).

An interactive tool for exploring the relationship between input variables and model outputs for each of the three stages of an outbreak is available at https://phoebelu.shinyapps.io/DepopulationModels/.

### 3.3 Impact of preemptive control by vaccination or depopulation

When our models of disease introduction, outbreak probability and outbreak size are combined, there is a significant impact of interventions that decrease the size of the susceptible population (Figure 3). Given that there is likely substantial variability in the value of the reproduction number within different congregate settings, it is notable that control interventions have a large impact for a wide array of *R*_0_ values. For example, when the susceptible population is decreased by 20% and transmission decreases linearly with control (i.e. *γ* = 0.2, and Θ = 1), the expected number of total infections decreases by 73%, 47%, 40%, and 38%, for *R*_0_ of 1.5, 3.0, 5.0, and 8.0 respectively (Bottom panel of Figure 3). These relative proportions hold for a broad range of values for other model parameters (data not shown).

**Figure 3.**
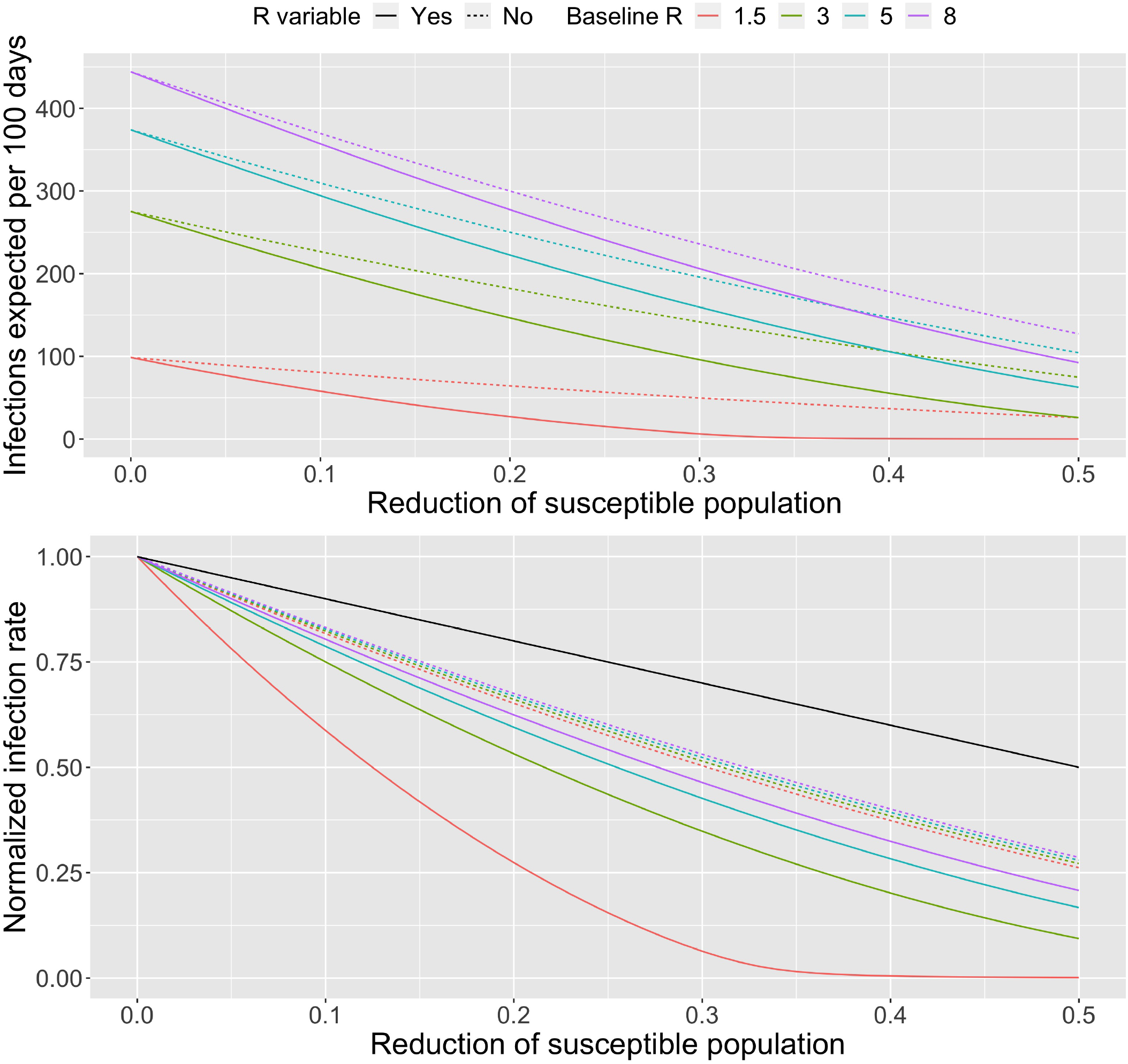
Impact of control measures that decrease the size of the susceptible population. (Top) The expected number of infections due to outbreaks occurring within 100 days as a function of the level of control. The control value, *gamma*, shown on the x-axis is the proportion of the number of susceptible individuals that are removed from the resident population via vaccination and/or depopulation. Colors correspond to different values of the baseline reproduction number, *R*_0_, for when no control is implemented. Specific values of the *R*_0_ are specified by the legend. Dashed lines show the result assuming that the control intervention does not change the reproduction number (e.g. Θ = 0 corresponding to a vaccine that immunizes against disease, but not asymptomatic infection). Solid lines show results assuming that the reproduction number changes in proportion to the level of control (e.g. Θ = 1 corresponding to a vaccine that immunizes against disease and asymptomatic infection). (Bottom) Analogous to top panel, except that the rate of infection has been normalized to a rate of one when the level of control is zero. This highlights the relative impact of decreasing the number of susceptible individuals. For both panels superspreading is allowed, which is modeled by a dispersion value of 0.2. We set *N*_*C*_ = 10 and *α*_*ic*_ = 0.01, meaning that residents are assumed to have 10 contacts with staff a day and that contact with an infected staff has a one percent chance of transmitting infection. The other parameters are the same as the defaults for Figures S1 – S3.

Notably, the reduction in the number of cases as a function of control occurs in greater than linear fashion (seen by colored lines falling below the black line in Figure 3). The decrease is even more significant if control impacts both the susceptibility to infection and the transmission potential of individuals, as compared to control affecting susceptibility alone (seen by solid lines falling below the dashed lines in Figure 3). The greater than linear impact of control can be explained by how the expected number of infections is the product of the probability of an introduction, the probability that an introduction leads to an outbreak and the size of an outbreak. All of these factors depends on either the effective reproduction number, the size of the susceptible population, or both. Since control decreases the size of the susceptible population and possibly the effective reproduction number as well, each stage of outbreak dynamics can contribute to the reduction in the number of cases when control is applied (Figures 1 and S4). When the dependency of these individual components of outbreak dynamics are multiplied together, the result is a greater than linear impact of control. The extreme values of the susceptibility index considered here (i.e. Θ = 0 and Θ = 1) provide bounds for the expected dynamics of intermediate values.

If control reduces the reproduction number linearly (i.e. Θ = 1), our model predicts several changes in the dynamics of outbreaks as control is applied (Figure S5). Notably, the attack rate amongst susceptible individuals decreases as the effective reproduction number, *R*, decreases (Figure S5, upper right panel), particularly as *R*_0_ approaches one. This implies that for relatively small *R*_0_, the decrease in the size of the outbreak seen with control is largely driven by the attack rate rapidly approaching zero, rather than the size of the susceptible population itself. However with higher values of *R*_0_, most of the decrease in the size of the outbreak is driven by a decrease in the number of susceptible residents rather than by a reduction in the attack rate. Reducing the size of the susceptible population also delays the timing of the peak for the number of infectious individuals and reduces the maximum number of individuals infected at once (Figure S5, bottom panels). This can help facilitate roll-out of public health interventions that can further decrease the burden of disease. It also helps to minimize overcrowding of healthcare facilities used to treat severe cases. If control has no impact on the reproduction number (i.e. Θ = 0), our model predicts that the control has no impact on the attack rate or temporal dynamics of uncontrolled outbreaks.

### 3.4 Adjunctive strategies for disease control

In some circumstances, it may be possible to reduce the risk and burden of disease outbreaks by optimizing the way a group of residents are proportioned into distinct residential units, such as two independent buildings (Supplementary text S7.2.1). For most reasonable parameter values, it appears optimal to distribute residents in a manner that keeps *R* consistent across the population.

In other circumstances there may be an opportunity to implement reactive disease control measures in which the number of susceptible individuals is rapidly reduced once an outbreak has been detected to occur (Supplementary text S7.2.2). This contrasts with the preemptive approach discussed above of reducing the susceptible population before an outbreak has even occurred. When the efficacy of a reactive control is compared to preemptive control, it appears that reactive control is more likely to be effective when the reproduction number is lower, there is minimal delay in implementing control and the individual-level efficacy of reactive control is high.

## 4 Discussion

The burden of COVID-19 within prisons, nursing homes and other congregate settings has provided a devastating reminder of the fragility of congregate settings. To reduce unnecessary death, disease, and economic loss, policies are needed to prevent outbreaks from occurring and to mitigate outbreaks that have already started. Besides protecting the residents of congregate settings, interventions that reduce outbreak potential also reduce strain on local health systems and spillover infections in the community. An additional consideration is the strain on staff during outbreaks due to their own health risk, often being overworked, and the emotional burden of taking care of vulnerable populations. Thus the longer term implications of reducing outbreak potential include decreasing the risk of staff burnout, post-traumatic stress disorder, and loss of institutional trust.

Our model for the impact of control on outbreak dynamics can be integrated into a variety of infection control policy decisions. For example, in the context of the COVID-19 pandemic, our study of the impact of decreasing the susceptible population could have provided quantitative context to decisions about how much to prioritize decarceration of prison populations, or whether additional nursing homes for COVID-19 patients are needed to ensure that existing ones are not overburdened with transmission. In particular, we found that relatively small reductions in the susceptible population of congregate settings could have a significant impact on the overall number of cases due to outbreaks. This multiplicative effect occurs due to a combination of decreasing the rate of disease introductions, the probability that an introduction leads to an outbreak and the size of any outbreak that occurs (Figure 1). Consideration of how populations can be divided into distinct residential units as well as the possibility of reactive interventions once an outbreak begins provides further opportunities for disease control.

In order to frame our analysis of control strategies in a manner that was intuitive and transparent, we made many assumptions. Depending on specific circumstances of spread in congregate settings, some assumptions will be more relevant than others. Specifically we ignore the possibility of direct transfer of infected individuals from one building to another as can inadvertently happen for individuals who are transferred during the latent period. We ignore the fluctuation of disease prevalence in the staff, particularly as surges may occur due to staff-staff transmission, resident-to-staff transmission, or increases in community transmission. Similarly we ignore how staff-resident contacts may evolve over the course of a pandemic, particularly as access to personal protective equipment and education about infection control improves over time. Our use of a SEIR transmission model assumes that within a residential unit (e.g. an isolated building) everyone is in equal contact with each other and thus ignores the finer scale structure of population dynamics. Our SEIR model also incorporates the standard assumptions of compartmental models including constant exponential rates of transitions and standard mass-action transmission. These assumptions ignore the possibility of seasonal effects, such as the possibility that transmission is impacted by changes in air circulation when heating or air conditioning is utilized. Except for our modification for reactive vaccination, The SEIR model also ignores how a large outbreak would likely inspire multiple efforts to acutely mitigate disease transmission. Finally, our assumption of a linear, density-dependent relationship between the size of the susceptible population and the reproduction number ignores how factors such as the relationship between duration-of-exposure and infection status may yield a non-linear relationship.

Our model can be adjusted to accommodate more flexible assumptions, but this would come at the cost of adding extra parameters whose values may not be readily identifiable. Importantly, the key finding that decreasing the number of susceptible individuals in congregate settings can have a greater than linear impact on disease burden is expected to be robust to many variants of our model. It is also important to place interventions that reduce the susceptible population into a large context of intervention strategies. Many scenarios may benefit from other control interventions such as improved ventilation, more frequent testing, greater adherence to social distancing, improved utilization of personal protective equipment, or cohorting of staff by residential unit. In addition, besides absolute case counts there are other important considerations for describing the impact of outbreaks on residents of congregate settings including contributions to health inequity, mental health toll, and reduction of ancillary services that are deprioritized during pandemic emergencies. Specific policy-making decisions also need to consider how control strategies will impact the ability of staff to fulfill their normal professional responsibilities, as well as protect their own health.

## 5 Conclusion

Congregate settings pose a risk of large disease outbreaks. To reduce the burden that outbreaks have on residents, staff and the community at large, it is important to optimize strategies for preventing and mitigating outbreaks. We find that preemptive reduction of the size of the susceptible population via depopulation or vaccination can have a greater than linear affect on outcomes.

## Data Availability

All data sources are publicly available, as indicated in the methods section

https://github.com/uclalawcovid19behindbars/data

https://phoebelu.shinyapps.io/DepopulationModels/

https://github.com/proctor-ucsf/Transmission-in-congregate-settings

## 7 Supplement

### 7.1 Supplement text: Methods

#### 7.1.1 Probability of introduction

We model introduction of disease into the resident community as primarily coming from contact with infected staff. The daily rate of introducing a case, *ϕ*, is the product of the average number of staff-resident contacts per day and the probability that a staff-resident contact causes transmission. The average number of staff-resident contacts per day is the product of the number of susceptible individuals in the resident community, *N*_*s*_, and the average number of contacts residents have with staff, *N*_*c*_. Note that we define a contact to be a pairing of a resident and staff member. If there are multiple interactions of that pairing through a day, we still count it as one contact. We also ignore staff contacts with residents who are not susceptible as these will not result in disease introductions. The probability that a staff-resident contact causes transmission is the product of the prevalence of disease in the staff, *P*_*com*_, and the probability that an infectious contact causes an infection *α*_*ic*_,. Thus,

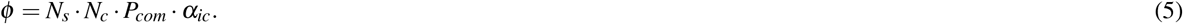

The average number of days until an an introduction occurs is the reciprocal of *ϕ*. If there is significant concern for introduction from visitors, then this can be accommodated by defining *N*_*c*_ as the sum of average daily contacts a resident has with both staff and visitors. The various components of Equation 5 are likely to change over time. For the purposes of evaluating the impact of interventions that decrease the size of the susceptible population, we consider *ϕ* to be an average daily rate of introducing a case.

#### 7.1.2 Probability of an introduction going extinct

When the number of residents with infection is a low number, the stochasticity of transmission can lead to significantly different outcomes. In some cases, introduction of disease may lead to no additional infections or a limited number of infections. In other cases, introduction of disease may lead to enough cases such that an outbreak is inevitable. We refer to the probability that an outbreak occurs as the probability of uncontrolled transmission, *p*_*uc*_.

To model the probability that introduction of disease leads to extinction, we assume that the probability distribution for the number of secondary infections caused by each new infection follows a negative binomial distribution.^26^ The negative binomial distribution is described by the reproduction number, *R*, and the dispersion parameter, *k*. The reproduction number is the average number of cases each new infection causes, but differs from *R*_0_ in that it incorporates possible reduction of the susceptible population due to vaccination or depopulation. The dispersion parameter characterizes the degree of transmission heterogeneity. Low values of *k* are seen when superspreading occurs, meaning that a relatively few number of cases causes a large proportion of the cases.

With these assumptions, there is an analytic relationship between *R, k*, the number of disease introductions occurring concurrently, *ζ*, and the probability, *q*_*s*_(*ζ*), of having an infection cluster of size of *s*.^30,31^ The probability, *r*_*i, j*_, that *i* infections causes *j* infections is,

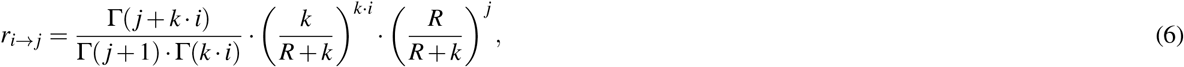

where Γ is the Gamma function. Then,

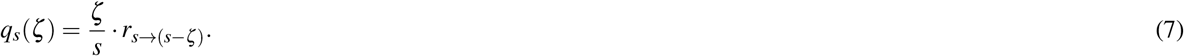

The use of a stationary negative binomial offspring distribution assumes that depletion of susceptibles is not a significant factor in transmission dynamics. This is a reasonable assumption if the number of infections that follow one or more introductions is small. However, once there are a significant number of infections following one or more introductions, a different transmission model is needed that accounts for depletion of susceptibles when there is uncontrolled transmission. We classify those instances in which there are more than *C*_*th*_ cases as uncontrolled transmission. All clusters of infection arising from an introduction that have less than *C*_*th*_ cases are considered extinction events. Thus the probability of *ζ* concurrent disease introductions leading to uncontrolled transmission is,

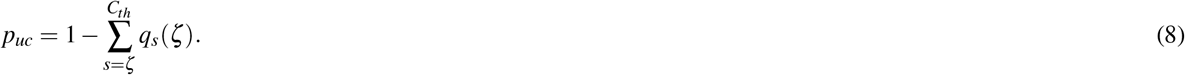

#### 7.1.3 Size and temporal dynamics of an outbreak

Once an outbreak is established, we use a deterministic compartmental model to describe the transmission dynamics. In this model, the rate that the susceptible residents become infected is proportional to the product of the number of susceptible and infectious residents. Once infected, a resident will be classified as exposed, but not infectious. The rate that exposed residents transition into being infectious is constant. Infectious residents are removed from the population at a constant rate. Removal may occur due to recovery, transport to a higher level of care or death. Once residents are removed, they are considered non-infectious and are no longer susceptible. During the course of an outbreak, it is assumed that no new susceptible residents are introduced into the population The equations describing the model are,

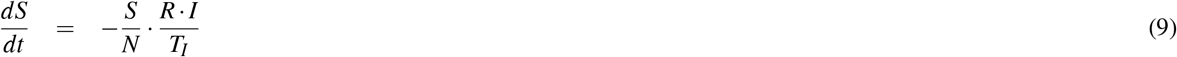

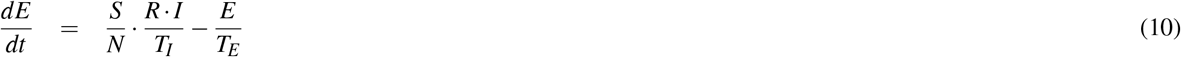

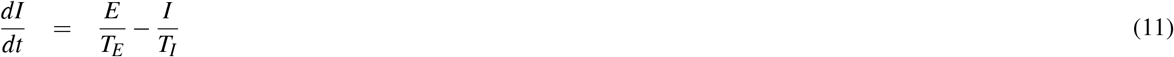

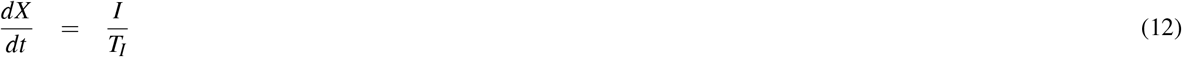

where *S, E, I*, and *X* represent the number of susceptible, exposed, infectious, and removed residents. The average time for being in the exposed and infectious states is represented by *T*_*E*_ and *T*_*I*_ respectively.

To determine the total number of residents infected, *I*_*tot*_, the proportion of residents who become infected, *P*_*T*_, the maximum number of residents infected at once, *M*_*T*_, and the time of peak incidence, *T*_*M*_, we simulate the preceding model numerically. Then,

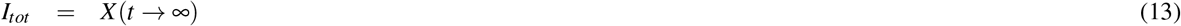

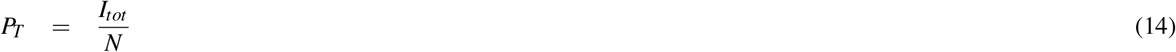

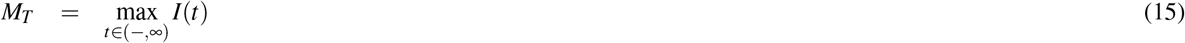

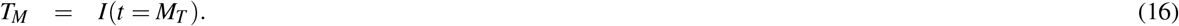

*P*_*T*_ is also known as the attack rate

For the simple compartmental model we are utilizing, other methods are available for determining the attack rate, such as transcendental equation relationships for the final size of epidemics.^32^ The benefit of utilizing a computational approach is that it offers a more flexible framework for making modifications to the model such as might occur if there were different risk classes or other forms of popuation structure.

#### 7.1.4 Optimizing distribution of residents

When considering the optimal proportion of residents to house in each of two independent residential units, we specify the baseline occupancy of each residential unit, 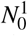 and 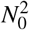, as well as the basic reproduction numbers at those baseline occupancies assuming all residents are susceptible, 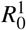 and 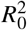. We also specify the fraction of population that are immune due to vaccination or prior infection, *V*. For a proportion *σ* of residents assigned to residential unit one, we set the size of the susceptible pool and the effective reproduction numbers prior to any outbreaks as,

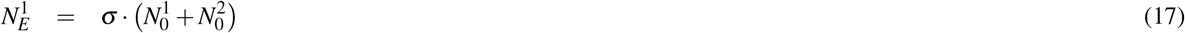

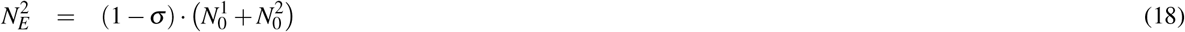

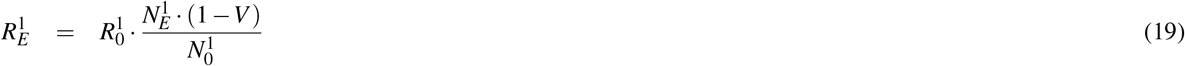

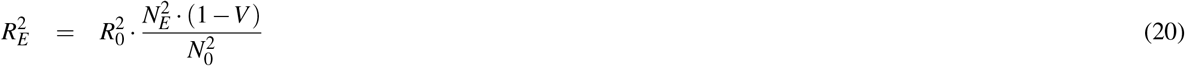

Except for these parameterizations, the model is run for each residential unit independently to determine the outbreak probabilities for each unit, 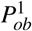 and 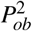, and expected outbreak sizes, 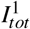 and 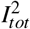. The joint probability of an outbreak occurring in at least one residential unit and the overall expected number of cases due to outbreak dynamics is then,

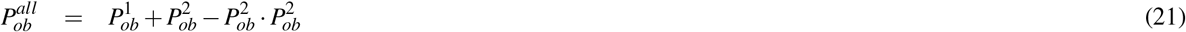

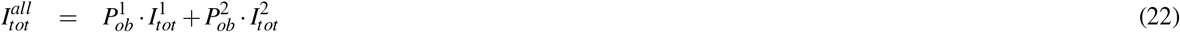

#### 7.1.5 Reactive control

As an alternate to preemptively vaccinating residents before an outbreak occurs, we also consider a reactive approach. In this scenario, we allow the baseline model to run until a ‘trigger threshold’ is reached for the number of cases and then vaccination is provided to the residents in the affected unit. The model continues to run unchanged for a time period specified by the control delay. Then, the susceptible population experiences a one time relative decrease of one minus the reactive vaccine efficacy. The reactive control efficacy is distinct from the ‘preemptive vaccine efficacy’ denoted by *V* (Section 7.1.4). The rest of the model proceeds as before.

### 7.2 Supplement text: Results

#### 7.2.1 Optimizing distribution of residents

Decreasing the size of the susceptible population is not the only way to decrease the probability of an outbreak or the expected number of cases from an outbreak. These quantities can also be reduced by cohorting residents between two independent residential units. Independent residential units might be different buildings within a prison, different units in a healthcare facility, or other housing paradigms that exclude mixing of residents from different units. The optimal distribution of susceptible residents between two separate residential units depends on the relative transmissibility in each unit, and the overall number of susceptible residents (Figure S6).^33^ As described in the supplemental methods, the transmissibility is characterized by the reproduction number corresponding to baseline occupancy by an entirely susceptible population of residents.

To minimize the probability that an outbreak occurs, the optimal distribution of susceptible residents is the one that equalizes the reproduction number between the residential units (Figure S6, top). Thus if the residential units have the same transmission potential, the outbreak probability is minimized by keeping the residential unit occupancy balanced at 50% each. However in Scenario 2 where the reproduction number for Unit A and B is 6 and 2 respectively, then the outbreak probability is minimized when 25% of residents are in residential unit A and 75% are in residential unit B. At this occupancy, the reproduction number equal in both residential units becomes three (Section 7.1.4).

The optimal proportion of residents to house in each residential unit to minimize the expected number of cases from outbreaks does not follow a simple rule (Figure S6, bottom). For scenarios with moderate values for the probability of an outbreak occurring, the optimal proportion to house in each residential unit to minimize the expected number of cases is closer to 50% than what is expected from analyzing the outbreak probability alone. For scenarios with low values of the probability of an outbreak occurring, the expected number of cases due to outbreaks occurring can remain low for a wide range of the proportion housed in each residential unit (Scenario 4). This is because the effective reproduction number can be kept below one across this range and thus outbreaks are not expected to occur. For high risk scenarios where the probability of an outbreak occurring is close to one, the best opportunity to minimize the number of cases is to reduce the number of residents in one residential unit to the point that a large outbreak can only occur in the more populous residential unit (Scenario 6). However, the benefit of the optimal distribution in the high risk scenario is quite marginal. The intricacies of these relationships can be visualized by considering the outbreak dynamics in each residential unit separately (Figure S7).

#### 7.2.2 Preemptive versus reactive control

When resources are constrained, there can be a trade-off between preemptive and reactive strategies for outbreak control. For example, suppose there are five equally sized residential units and only enough vaccine for 20% of the total population. As a preemptive approach, the number of susceptible residents in each residential unit could be reduced by 20%. An alternative reactive approach would be to monitor for disease introduction and if evidence of within-residence transmission is detected, then vaccinate all residents in the residential unit as quickly as possible.

Visualization of the outcomes for preemptive vs reactive vaccination shows that the benefit of a reactive strategy depends heavily on the control delay and the effectiveness of vaccination (Figure S8). Reactive control is more likely to be superior to preemptive control if the reproduction number is low. The higher the reproduction number is, the quicker a reactive strategy will need to be deployed in order for it to have much impact. Even with a control delay of 30 days our model predicts a reactive vaccine efficacy of 20% would be superior to preemptive vaccine efficacy of 10% for an *R*_0_ of 1.5 (seen by the red line being below the dotted line in the left panel of Figure S8). However, if *R*_0_ were 3.5, even rapid implementation of reactive control with an efficacy of 20% would not yield better results than preemptive control with an efficacy of 10% (seen by the red line being above the dotted line in the right panel of Figure S8).

Our definition of preemptive vaccine efficacy combines the impact of preemptive vaccine coverage and the ability of the vaccine to prevent transmission. For example, if two third of residents accept the vaccine and the vaccine decreases transmission by 75% then the preemptive vaccine efficacy is 50%. The reactive vaccine efficacy is meant to incorporate a combination of effects including imperfect vaccination rates, delays in vaccine-induced immunity and breakthrough infections despite maximum vaccine immunity. One caveat of these results is that we have assumed that there is always enough vaccine available to implement reactive vaccination. If there are outbreaks in multiple residential units that require a reactive response, then the reactive vaccine efficacy may decrease substantially due to vaccine shortage.

Besides reactive vaccination, it may also be possible to implement a reactive depopulation strategy. However, it would be important that the residents who are removed from the outbreak are quarantined prior to moving to a new location. Otherwise reactive depopulation could seed outbreaks in new locations.

## 8 Supplemental Figures

**Figure S1.**
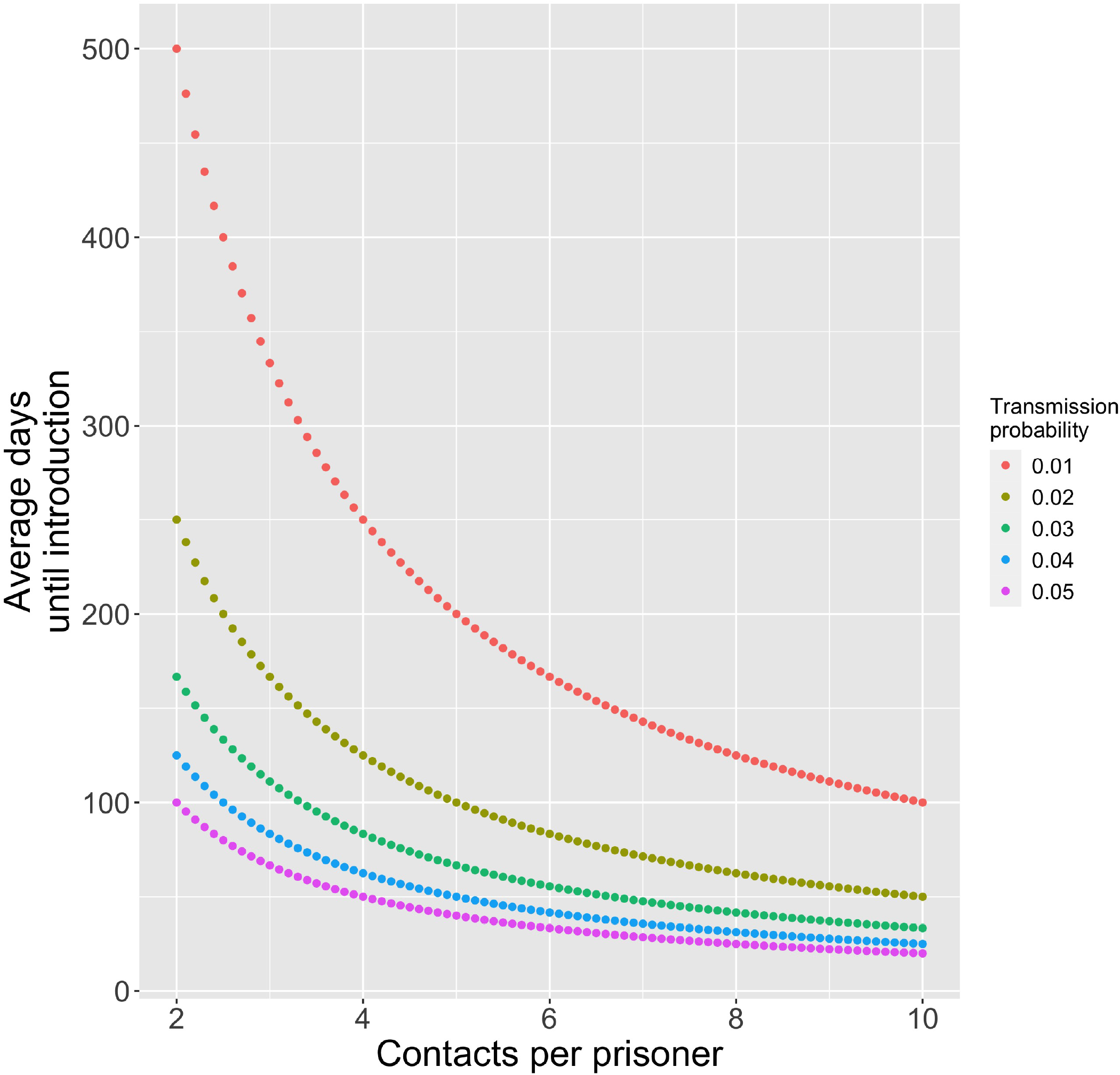
Frequency of an introduction. Average number of days until a COVID-19 case is introduced into the resident community, *ϕ*, as a function of the average number of contacts a resident has with resident staff, *N*_*c*_. Colors correspond to different values for *α*_*ic*_, the probability that a resident’s contact with an infected staff causes an infection. The prevalence of disease in the community, *P*_*com*_, is assumed to be 0.01%. The number of susceptible individuals in the congregate community, *N*_*s*_, is assumed to be 1,000.

**Figure S2.**
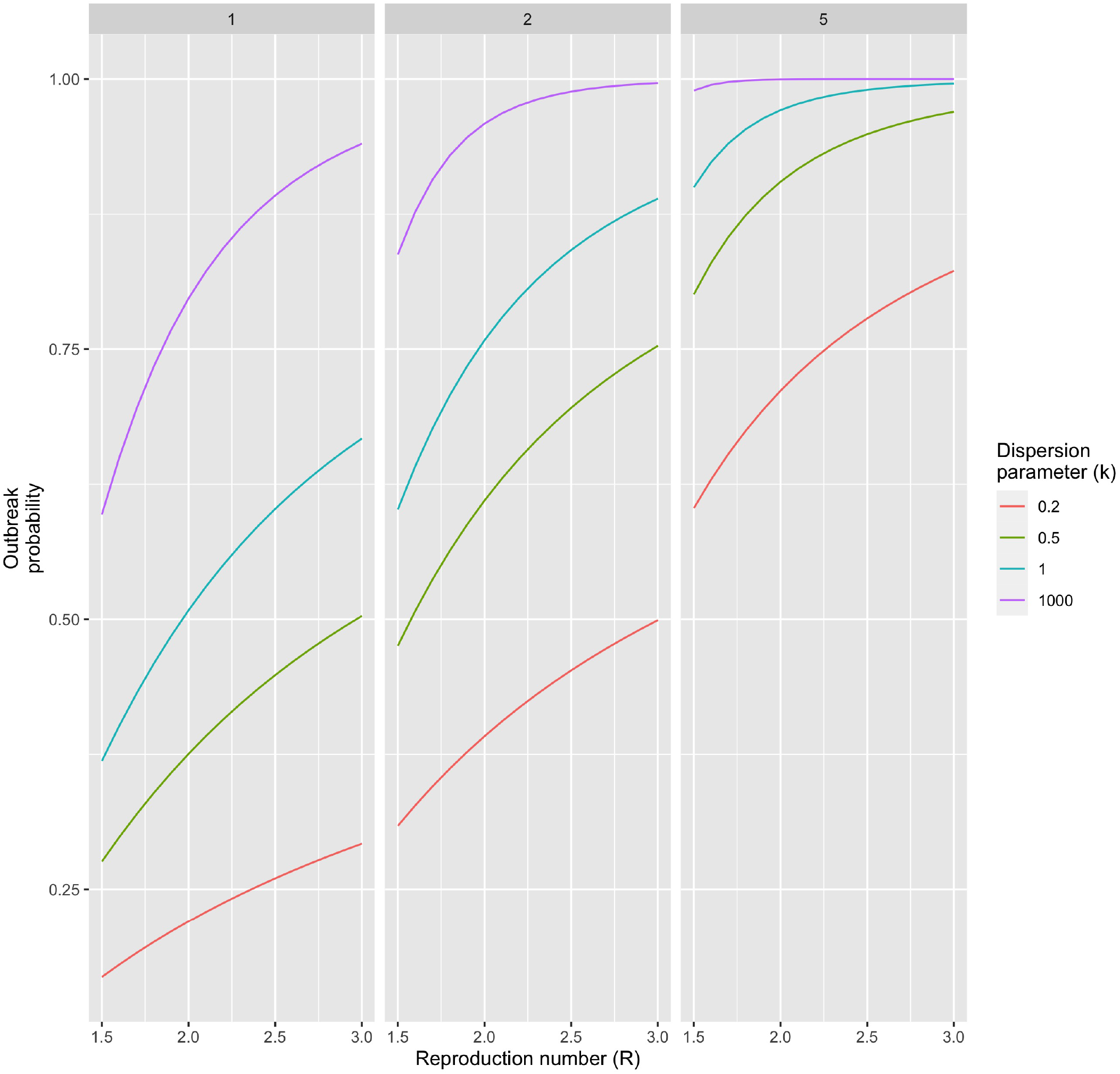
Probability of an outbreak. The probability of an outbreak occurring as a function of the reproduction number, which is defined the average number of transmission events each new case causes. The panels correspond to different numbers of simultaneous disease introductions as indicated in the top of the panel. The different colors correspond to different values of the dispersion parameter. Homogeneous transmission corresponds to *k* = ∞, and superspreading is more prevalent as *k* decreases. Plots are based on *C*_*th*_ = 10, meaning that an outbreak is defined to occur when an introduction leads to at least ten cases.

**Figure S3.**
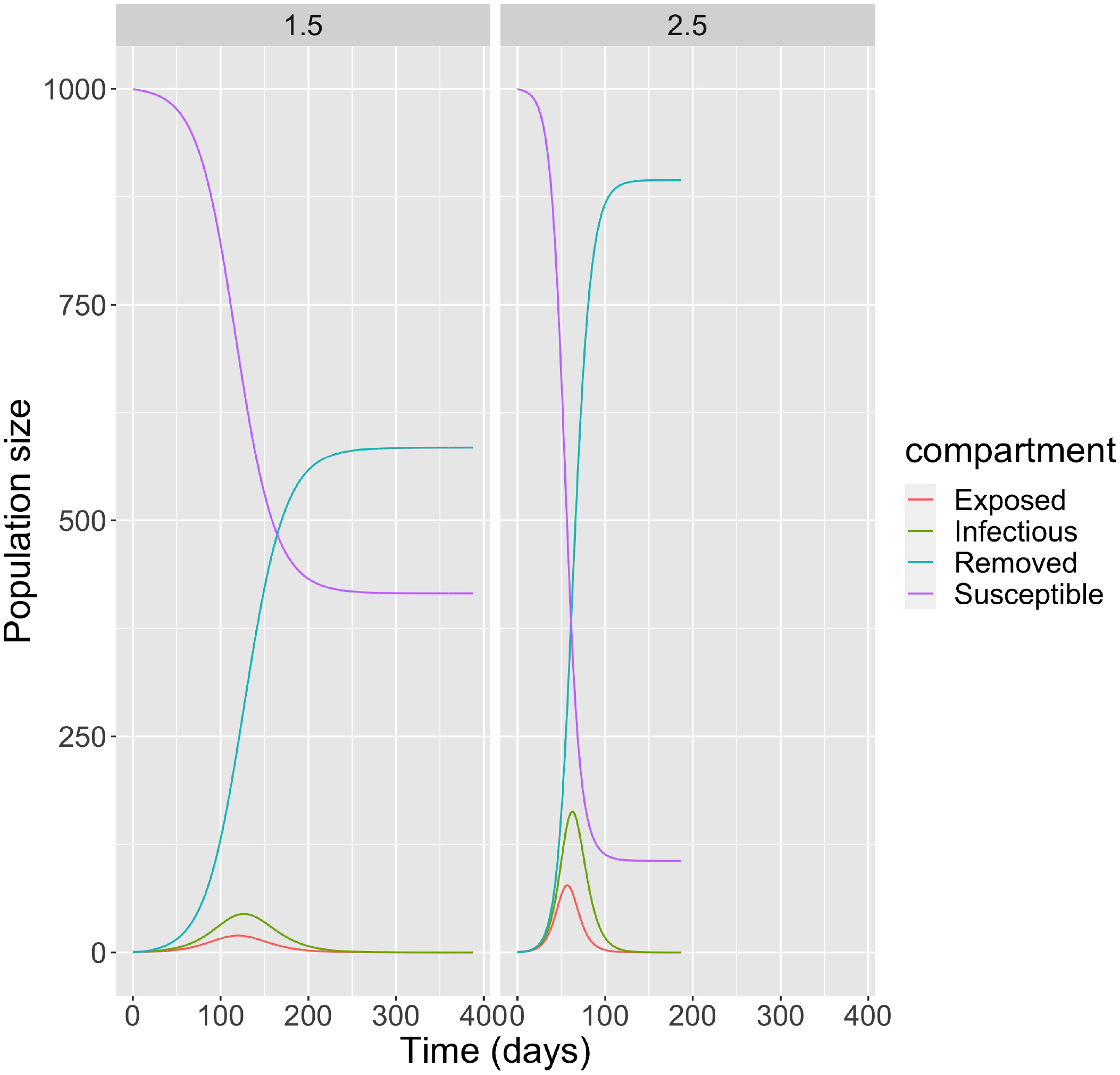
Transmission dynamics. The number of susceptible, exposed, infectious and removed residents are shown as a function of time for an outbreak in a congregate setting. The removed category includes those who have recovered from illness, those who are sick but quarantined and those who have died. The two panels represent two different values of R A total population of 1,000 is assumed. The average duration of each COVID-19 case being in the latent and infectious periods is 3 and 7 days respectively. The depicted outbreaks start with one exposed individual at time 0. The time step used for running the transmission dynamics model is 0.2 days, and the model is run until a negligible number of infectious individuals remain.

**Figure S4.**
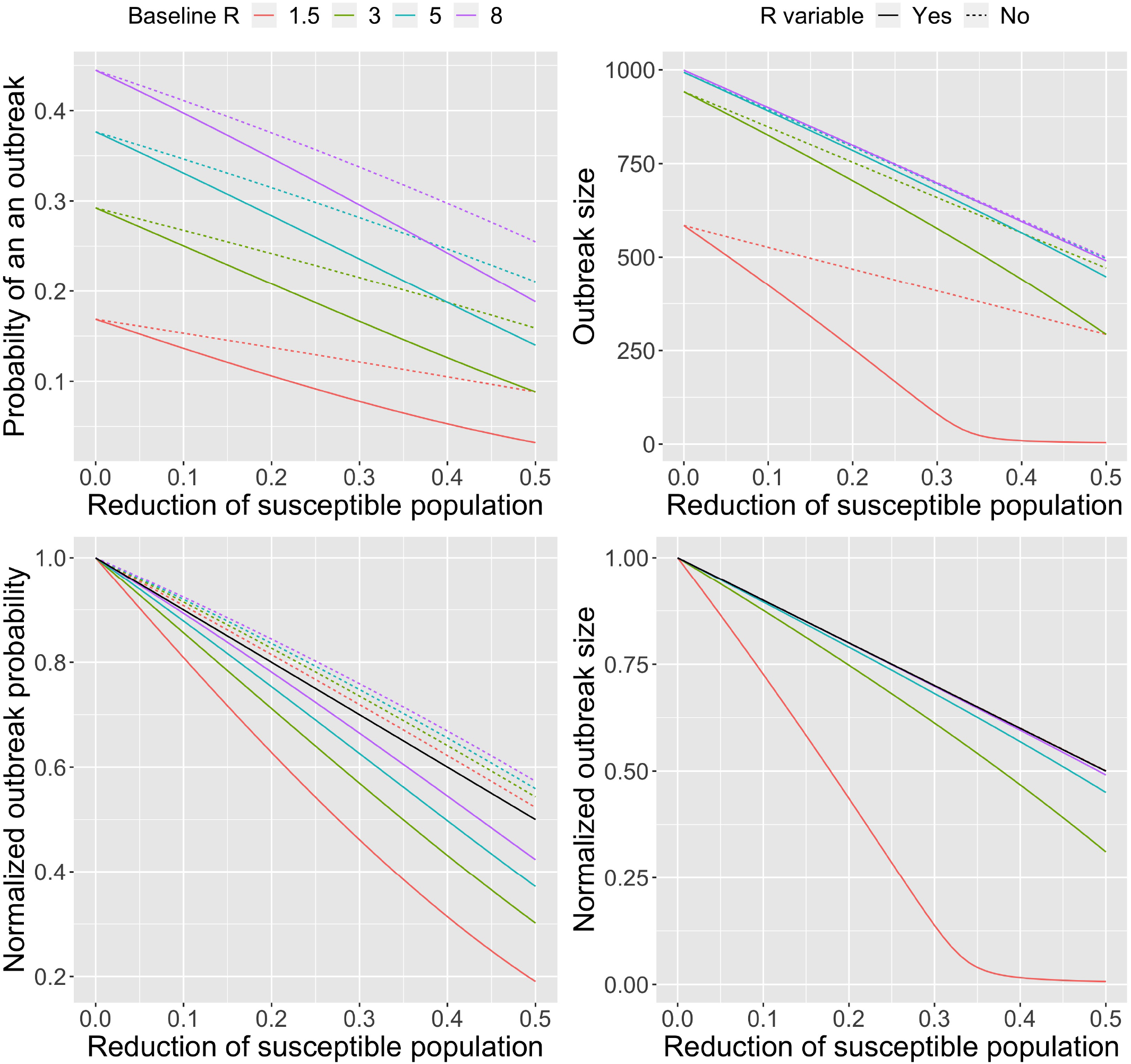
Probability and size of an outbreak. Probability of an outbreak occurring in 100 days (left panels) and size of outbreaks (right panels) as a function of control. The level of control is defined as the proportion of the susceptible population reduced by processes such as vaccination or depopulation. As parameterized by the legend, colors correspond to different values of the baseline reproduction number that represents transmission when there is no control. Dashed colored lines represent the modeling assumption that the reproduction number is unaffected by control (i.e. Θ = 0). Solid colored lines represent the modeling assumption that the reproduction number varies in direct proportion to the fraction of the population that remains susceptible (i.e. Θ = 1). Top panels show the absolute values, while the bottom panels show normalized values that are always one when there is no control. The black line in the bottom panel is a visual aid that signifies how the outbreak probability and outbreak size would change if there was a linear relationship with the level of control. Parameter values are as specified in Figure 3. As a technical note, there is a less than linear impact on the probability of an outbreak if control does not affect the reproduction number (dashed lines in lower left panel), because for our chosen parameter values the probability of an outbreak begins to saturate when the level of control is very small.

**Figure S5.**
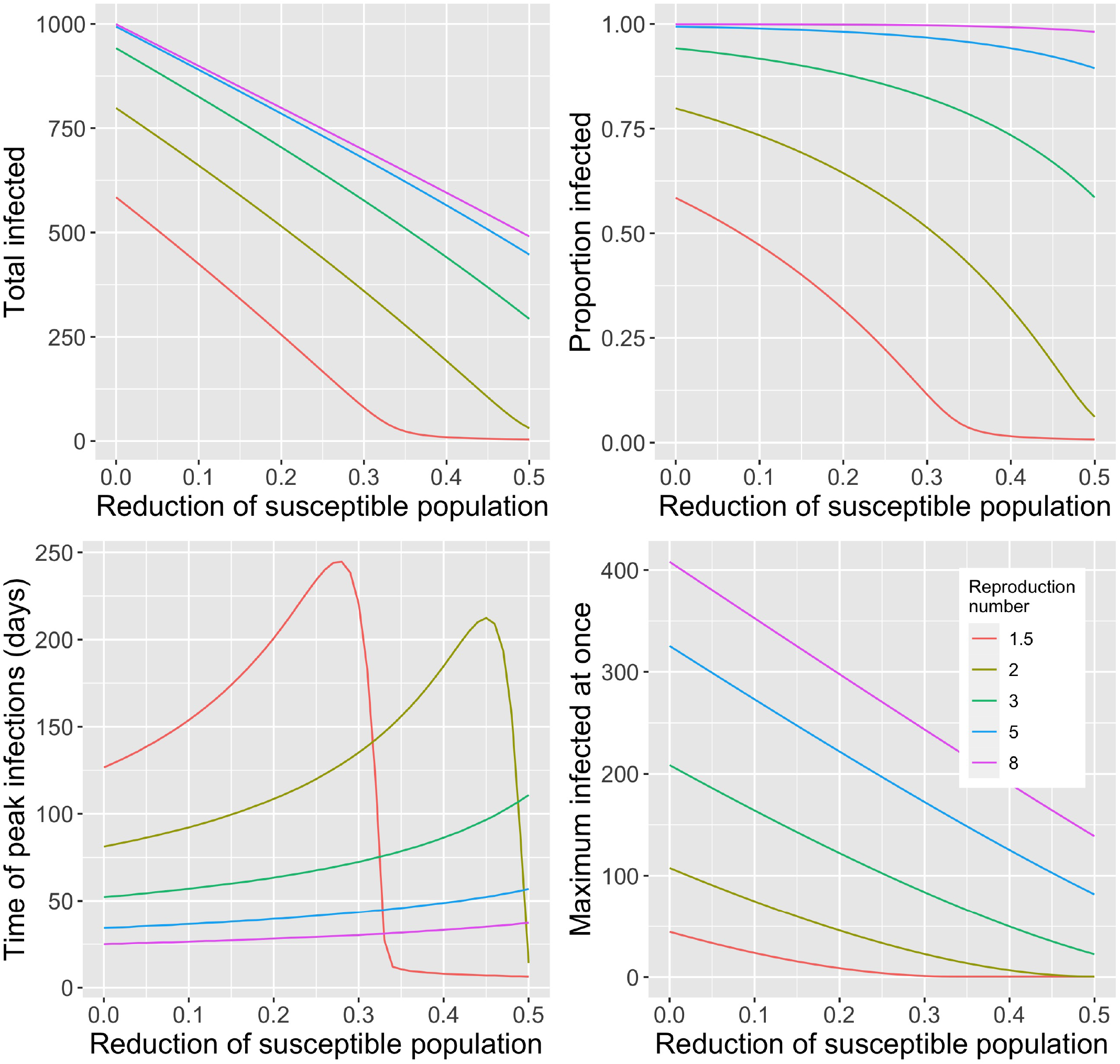
Outbreak size and timing. Each panel shows a feature of outbreak dynamics in a single residential unit as a function of control, which we have defined as the proportion of the population that is no longer susceptible due to processes such as depopulation or vaccination. The different colored lines correspond to different values of *R*_0_, which is the reproduction number when there is no decrease in the size of the susceptible population. Results are shown for the assumption that the effective reproduction number scales directly with proportion of the population that remains susceptible (i.e. Θ = 1). The panels show the impact on the total number of residents who are infected (upper left), the proportion of residents who were susceptible at the start of the outbreak that become infected (upper right), the timing of the peak in the number of infectious individuals (lower left), and the maximum number who are infectious at once (lower right). Parameter values are the same as for figure S3. As a technical note, for lower values of baseline *R*_0_ the timing of the peak in the number of infectious individuals abruptly decreases when the control factor is sufficiently high because the effective reproduction number becomes less than one at the start of an outbreak and so no significant peak in infectious cases occurs.

**Figure S6.**
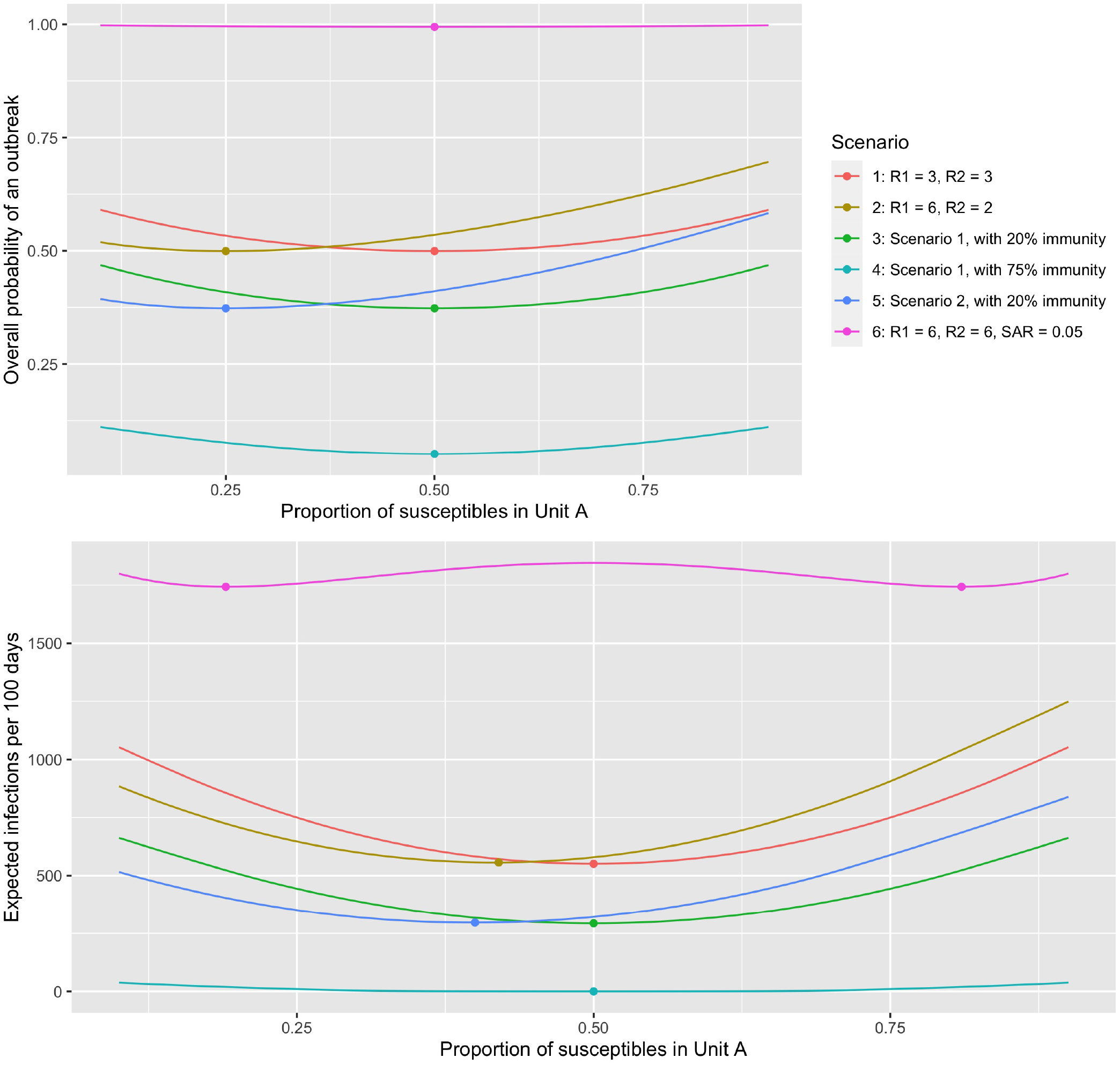
Optimal distribution of residents between two independent residential units. (Top) Probability that an outbreak occurs in at least one of two residential units within 100 days. We denote the residential units as ‘Unit A’ and ‘Unit B’. Results are shown as a proportion of the susceptible population that is housed in Unit A. The total number of residents is assumed to be 2,000 so that if 20% of the population have immunity from vaccination, a proportion of residents housed in Unit A of 0.25 corresponds to 400 susceptible residents in Unit A and 1,200 in Unit B. The colored lines represent six different scenarios as indicated by the legend. The reproduction values in the legend correspond to the reproduction number if residents are all susceptible and distributed equally in both residential units. In scenario 1 the transmission in the two residential units are identical. In scenario 2, Unit A has higher transmission than Unit B. In scenario 3, which is designed to model a very risky housing environment, the probability that an infected staff transmits infection to a resident is increased five-fold compared to the other scenarios and the reproduction number for both residential units is high. Scenarios 4, 5 and 6, are identical to scenarios 1 or 2 except that a fraction of the residents in each residential unit have immunity from vaccination or prior infection. (Bottom) The expected number of infections per 100 days for both residential units combined is shown as a function of the proportion of residents in Unit A. The bottom panel is otherwise analogous to the top panel. Except as noted above, the parameter values are identical to figure 3.

**Figure S7.**
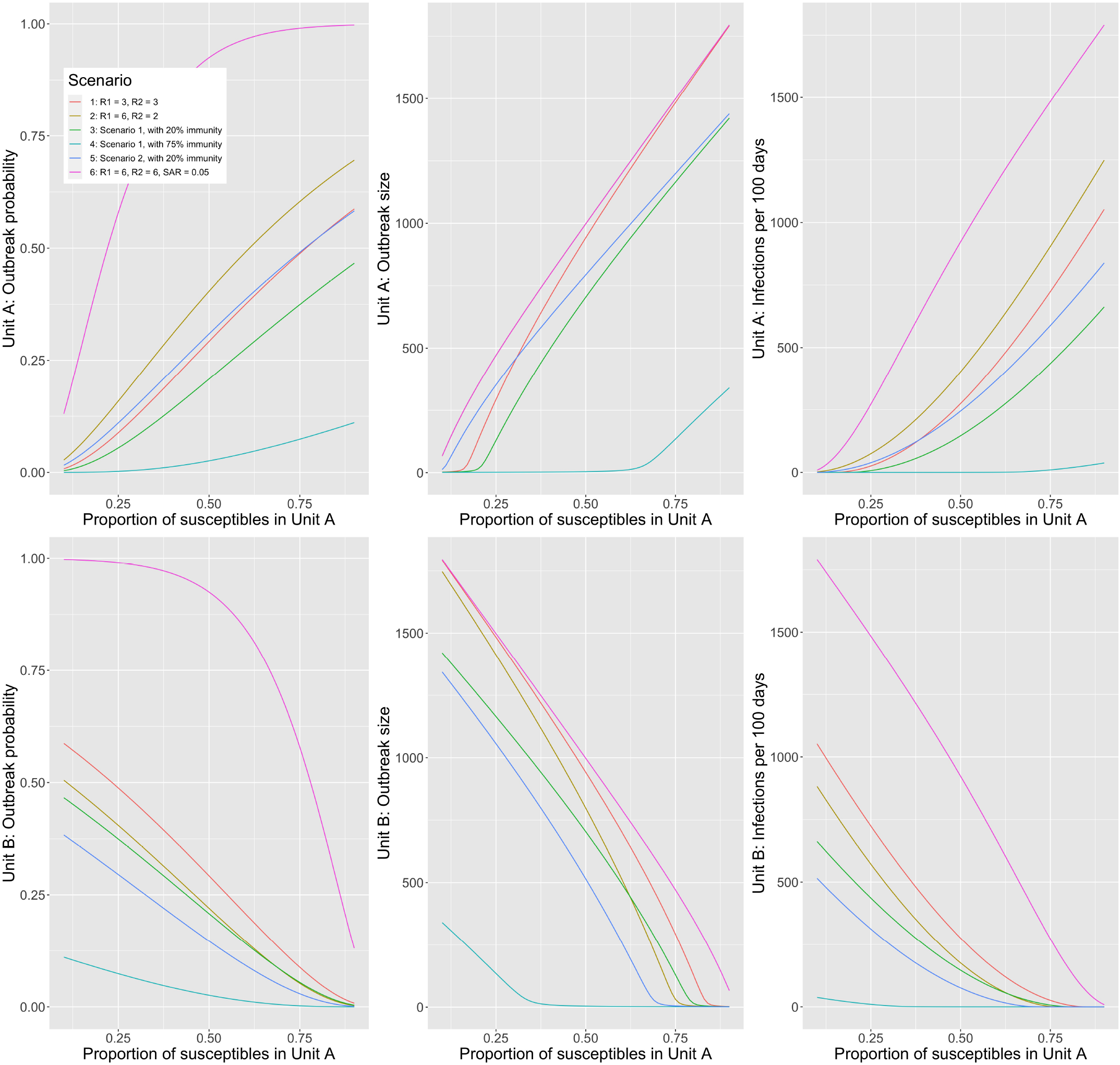
Outbreak dynamics depend on how residents are distributed between two residential units. The probability of an outbreak occurring in 100 days (left panels), expected outbreak size (middle panels) and the overall expected number of cases in 100 days (right panels) are shown as a function of the proportion of residents housed in Unit A. Top panels corresponds to Unit A and the bottom panels correspond to Unit B. Parameter values and scenarios are the same as for Figure S6.

**Figure S8.**
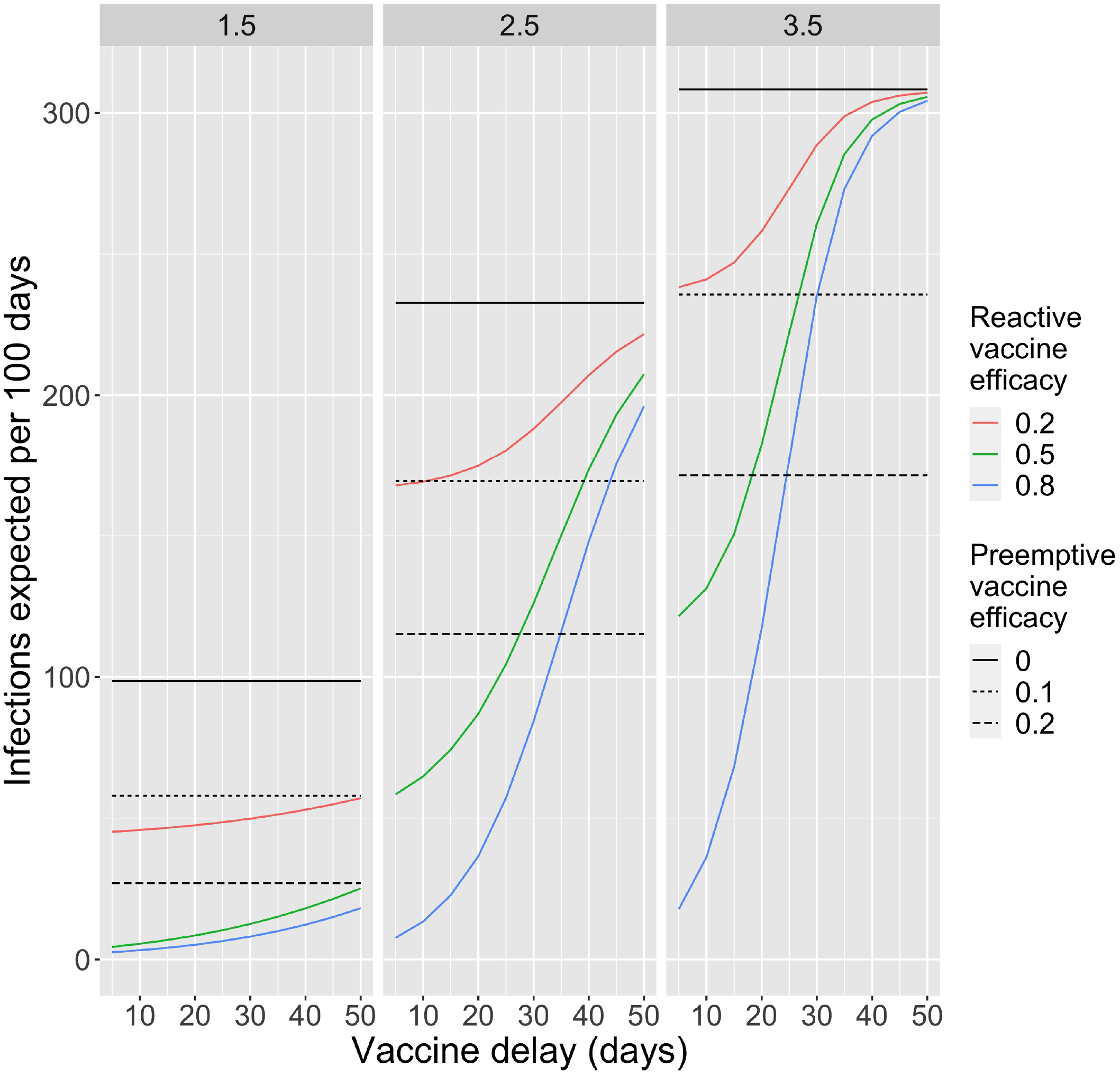
Preemptive vs reactive control. The number of infections per 100 days as a function of the vaccine delay for reactive vaccination. Each vertical panel corresponds to a different value of *R*_0_ as depicted on the top bar. Horizontal black bars correspond to no control being implemented. Horizontal dashed bars correspond to preemptive control, in which immunity from vaccination has been achieved before an outbreak begins. The individual-level efficacy of preemptive vaccination is indicated in the lower legend. Colored curves indicate the expected number of infections when reactive control is implemented once ten cases in a residential unit are identified. We model reactive control as being effective at reducing transmission after a predetermined vaccine delay. The delay is depicted on the x-axis and the individual-level reactive vaccine efficacy is depicted in the upper legend. Parameter values are otherwise identical to those in Figure 3.

